# Health-related quality of life in parents of long-term childhood cancer survivors: a report from the Swiss Childhood Cancer Survivor Study - Parents

**DOI:** 10.1101/2024.10.28.24316262

**Authors:** Sonja Kälin, Julia Baenziger, Luzius Mader, Erika Harju, Fabienne Gumy-Pause, Felix Niggli, Grit Sommer, Gisela Michel, Katharina Roser

**Affiliations:** Faculty of Health Sciences and Medicine, University of Lucerne, Lucerne, Switzerland; Cancer Registry Bern Solothurn, University of Bern, Bern, Switzerland; School of Health Sciences, ZHAW Zurich University of Applied Sciences, Winterthur, Switzerland; Department of Women, Child and Adolescent, Onco-Hematology Unit, Geneva University Hospitals, Geneva, Switzerland; University Children’s Hospital, University of Zurich, Switzerland; Institute of Social and Preventive Medicine, University of Bern, Bern, Switzerland

**Keywords:** health-related quality of life, parents, childhood cancer survivors, long-term

## Abstract

**Purpose:** Having a child with cancer can profoundly impact parents’ health-related quality of life (HRQOL). However, there is a lack of knowledge about the long-term effects of childhood cancer on parents’ well-being. The current study aimed to 1) describe the HRQOL of parents of long-term childhood cancer survivors (CCS) and compare it with that of parents from the general population in Switzerland, and 2) investigate sociodemographic and cancer-related determinants of lower HRQOL in parents of CCS.

**Methods:** In this cross-sectional study, a total of 751 parents of CCS (mean time since diagnosis = 23.7 years, SD = 6.7 years) and 454 parents from the general population reported their HRQOL by completing the Short Form-36 (SF-36v2). Sociodemographic and cancer-related characteristics were also collected.

**Results:** Multilevel regression analyses showed that parents of CCS and parents from the general population had similar physical and mental HRQOL. When comparing mothers and fathers separately, there were no differences between the samples, except for higher HRQOL in the domain of physical functioning in mothers of CCS. Cancer-related characteristics were not associated with HRQOL in parents of CCS. Several sociodemographic characteristics such as being female, being from the French or Italian-speaking part of Switzerland, having a lower education, having a chronic condition, and having a migration background were associated with lower HRQOL.

**Conclusion:** Parents of CCS are doing well a long time after their child’s cancer diagnosis. Nevertheless, tailored support should be provided for at-risk demographic groups.

**Plain English summary:** Having a child with cancer might have a profound impact on various aspects of parents’ well-being, including their quality of life. However, not much is known about how parents are doing very long after their child’s cancer diagnosis. Our study aimed to describe the quality of life of parents of childhood cancer survivors and compare it to that of parents from the general population. Furthermore, we sought to compare mothers and fathers separately. We conducted a questionnaire survey with 751 parents of childhood cancer survivors, on average 24 years after the diagnosis. Additionally, 454 parents from the general population completed a similar questionnaire. We found that parents of survivors reported a similar quality of life to parents from the general population. When comparing mothers and fathers separately, we found no differences between the samples, except for higher quality of life in the domain of physical functioning among mothers of survivors compared to mothers of the general population. We conclude that overall, parents of childhood cancer survivors are doing well long after their child’s cancer diagnosis.

## Introduction

A childhood cancer diagnosis is a profoundly disruptive event, impacting not only the life of the child with cancer, but also the lives of their parents. As a result, parents are at risk for low health-related quality of life (HRQOL) [1]. HRQOL refers to a multifaceted assessment of how well an individual functions and perceives well-being in the context of their physical, mental, and social health [2].

Parents of children with cancer may experience low HRQOL at different stages following the cancer diagnosis. During active treatment, parents of young children reported lower HRQOL compared to population norms, particularly in the HRQOL-domains vitality, mental health, role limitations due to emotional problems, and general health perception [1]. Time since diagnosis also plays an important role. In a longitudinal study from Australia with parents of seriously ill/injured children, parental quality of life (QOL) was reduced at the time of diagnosis and normalized seven months after diagnosis for both psychosocial and physical QOL [3]. Similarly, in a Dutch study including parents of children with leukemia, mental QOL gradually improved to values similar to the Dutch reference values from shortly after the diagnosis up to three years after the diagnosis [4]. However, other studies found lower HRQOL in both physical and mental HRQOL in parents of children with cancer compared to normative data, even after three to four years after diagnosis [5, 6]. A French study of parents of childhood leukemia survivors found that seven years after diagnosis, QOL was lower in the physical health and social relationships domains and higher in the psychological domain compared to a French reference population [7].

Overall HRQOL of parents of very long-term childhood cancer survivors (CCS) was similar (10 years after diagnosis) [8] or even slightly higher compared to the general population reference data (21 years after diagnosis) [9]. However, more parents of CCS reported mental health problems compared to normative data [8]. It is crucial to investigate whether low HRQOL persists in parents many years after their child’s cancer diagnosis. This will help to determine the continued need for parental support beyond the early stages of their child’s illness.

Previous research revealed differences in HRQOL between mothers and fathers of children with cancer. Mothers reported lower mental [6] and psychological [7] HRQOL than fathers. When comparing parents within the same family, mothers had lower levels in various HRQOL domains such as cognitive function, sleep, and vitality. Longitudinal data suggested that this discrepancy between mothers and fathers decreased over time [10]. In addition to gender, several other factors have been associated with HRQOL in parents of CCS. A high level of education was associated with higher physical [6] and psychological [7] QOL. Parents’ QOL improved over time after cancer diagnosis [5, 7]. Additionally, certain cancer-related characteristics, such as having received craniospinal radiation or having had a relapse, have been identified as risk factors for lower parental HRQOL[7]. Another study found that lower HRQOL in fathers of children with cancer was predicted by type of diagnosis (retinoblastoma) and higher treatment intensity [5]. However, an Australian study found no association between age, gender, diagnosis, treatment, number of late effects, or time since diagnosis and parental HRQOL [8].

Given the limited knowledge about the long-term well-being of parents of CCS, we aimed to 1) describe the HRQOL of parents of very long-term CCS and compare it to parents from the general population in Switzerland, and 2) examine sociodemographic and cancer-related characteristics for lower HRQOL in parents of CCS.

## Methods

### Study participants and procedure

#### Parents of CCS

This study is part of a project investigating psychosocial late outcomes in parents of long-term CCS (Swiss Childhood Cancer Survivor Study–Parents; SCCSS-Parents). The Childhood Cancer Registry (ChCR) is a nationwide, population-based registry for all Swiss residents diagnosed <20 years of age with a cancer diagnosis according to the International Classification of Childhood Cancer—Third Edition (ICCC-3) [11] since 1976 [12]. For this study, we included all children registered in the ChCR who were aged ≤16 years at time of diagnosis, who survived ≥5 years after diagnosis, and who were at least 20 years and alive at time of study.

Parents of CCS received an invitation letter and, two weeks later, two questionnaires - one for each parent to complete individually with a pre-paid return envelope. We reminded non-responders after about four weeks, and again after about two months. Data was collected between January 2017 and February 2018.

#### Comparison parents

A representative sample of Swiss households from the general population was randomly selected according to the distributions of age, gender, and language region (German/French/Italian) in Switzerland from the Swiss Federal Statistical Office (SFSO). Household members eligible to participate were between 18 and 75 years in 2015 [13]. Eligible individuals received a letter informing them of the study. Approximately two weeks later, they received a questionnaire accompanied by a letter and a pre-paid return envelope. Non-responders received reminders after about five weeks. Data was collected between May 2015 and June 2016. The comparison parent sample for this study comprised participants from the Swiss general population sample with at least one child aged ≥20 years at the time of the study.

In both samples, all study materials were available in German, French, and Italian to cover the three main language regions in Switzerland. There were two versions of the invitation letter. In one version, parents of CCS were addressed as parents of a child with cancer and comparison parents were told that they were serving as a comparison group for a study related to childhood cancer (cancer-related study aim). In the other version, individuals from the general population and parents of CCS were addressed as the general population to participate in a health and well-being study, and childhood cancer was not mentioned (well-being study aim). In the cancer-related study aim, parents of CCS were contacted by the former treating pediatric oncology center, all other individuals were contacted by the team at the University of Lucerne. The study aims communicated to the participants are not focus of this study, but differences were examined.

### Ethical approval

The SCCSS-Parents study was reviewed and approved by the “Ethikkommission Nordwest-und Zentralschweiz (EKNZ)” on 26 March 2015 (Reference: EKNZ 2015-075). All participants provided informed consent.

### Measures

#### Health-related quality of life

The Short Form-36 version 2 (SF-36v2) is a widely used tool for evaluating HRQOL across two categories: physical and mental HRQOL [14]. It measures eight domains via 36 items, with all but one item assigned to a specific health domain. The eight health domains address facets of physical and mental HRQOL: physical functioning (10 items), physical role functioning (4 items), bodily pain (2 items), general health perceptions (5 items), vitality (4 items), social role functioning (2 items), emotional role functioning (3 items), and mental health (5 items). The item assessing perceived change in health was not used [16, 17]. Two summary measure scores can be created from the eight health domain scores, one for physical HRQOL (physical component score; PCS) and one for mental HRQOL (mental component score; MCS). Higher scores imply better HRQOL. We used validated versions of the SF-36v2 in German, French, and Italian. Translations of the SF-36 have been shown to be culturally appropriate and comparable [13, 17, 18]. The SF-36v2 needs 5 to 10 minutes to be completed, has a high acceptability, and good psychometric properties in general populations, and populations with childhood cancer or other chronic diseases [13, 17–21].

The SF-36v2 questionnaire data was processed in line with the SF-36v2 manuals [16, 17]. Raw scores of the parents of CCS sample were transformed to p-scores (percentage scores ranging from 0 to 100) for the health domain scale scores and to T-scores using norm-based scoring according to normative data from the Swiss general population [13]. Cronbach’s alpha was considered good ranging from 0.76 (general health perceptions) to 0.94 (bodily pain) across the eight health domain scales in parents of CCS [22].

#### Sociodemographic characteristics

For parents of CCS, the following sociodemographic characteristics were collected in the questionnaire: gender, age at study (years), migration background, educational attainment (see below), employment status (employed/unemployed/retired), living in a partnership (yes/no), and having a chronic health condition (yes/no). For comparison parents, gender, age at study, nationality, and country of birth were derived from the SFSO, and educational attainment, employment status, living in a partnership, and having a chronic health condition from the questionnaire. Age at study was divided into four categories: 18-25 years, 26-40 years, 41-65 years, and 66-75 years. Participants were considered to have a migration background if they were not born in Switzerland, were not Swiss citizens, or had not been Swiss citizens at birth. The language region was defined according to the language of the questionnaire. The highest level of education attained was categorized into compulsory schooling/vocational training/upper secondary education/university education [23].

#### Cancer-related characteristics

Cancer-related characteristics of CCS were obtained from the ChCR and included: gender, age at diagnosis (years), diagnosis according to ICCC-3 [12], cancer treatment (coded hierarchically: surgery only/chemotherapy (may have had surgery)/radiotherapy (may have had surgery and/or chemotherapy)/stem cell transplantation (may have had surgery, chemotherapy, and/or radiotherapy)), time since diagnosis (years), and relapse (yes/no). Late effects of the CCS were self-reported in the parents’ questionnaire (yes/no). The information on their child’s late effects was only available for the group of parents of CCS who have been contacted as parents of CCS (i.e., for whom the cancer-related study aim was mentioned; n=454, 60.5% of participating parents of CCS) and not for the ones having been contacted as part of the general population of Switzerland (well-being study aim).

### Statistical analysis

Missing values in the SF-36v2 data, were imputed with the mean of the available scale items in cases where at least half of the scale items were available [15]. For all other variables, no imputations were made. All analyses were conducted using the Stata software [24]. Figures were generated using R [25].

For the sample description, we used descriptive statistics, and chi-square tests (categorical variables) and t tests (continuous variables) to compare characteristics of parents of CCS to comparison parents, and survivors with participating to non-participating parents. Parents of CCS and comparison parents were statistically significantly different regarding number of children, migration background, employment status, partnership, chronic condition or health problem, and study aim mentioned (Table 1). When comparing mothers and fathers separately between the samples, mentioning of study aim and number of children differed significantly in both groups. Additionally, mothers differed in terms of migration background, employment status, and chronic condition, whereas fathers differed in terms of partnership status. We adjusted for these variables in further analyses.

**Table 1.** Description of parents of childhood cancer survivors (CCS parents; n = 751) and parents of the general population (comparison parents; n = 454).

|  | Parents of childhood<br>cancer survivors<br>(CCS parents, n = 751) | Parents of the general<br>population<br>(comparison parents, n = 454) |  |
| --- | --- | --- | --- |
|  | n (%) | n (%) | p value<sup>c</sup> |
| *Study aim mentioned* |  |  | **<0.001** |
| Yes | 454 (60.5) | 197 (43.4) |  |
| No | 297 (39.5) | 257 (56.6) |  |
| **Socio-demographic characteristics** |  |  |  |
| *Sex* |  |  | 0.752 |
| Male | 309 (41.1) | 191 (42.1) |  |
| Female | 442 (58.9) | 263 (57.9) |  |
| *Age at study<sup>a</sup>* |  |  | 0.127 |
| 41-65 years | 521 (69.4) | 297 (65.4) |  |
| ≥66 years | 227 (30.2) | 157 (34.6) |  |
| Missing | 3 (0.4) | 0 (0.0) |  |
| *Number of children<sup>a</sup>* |  |  | **<0.001** |
| 1 child | 24 (3.2) | 72 (15.9) |  |
| 2 children | 333 (44.3) | 232 (51.1) |  |
| >2 children | 349 (46.5) | 150 (33.0) |  |
| Missing | 45 (6.0) | 0 (0.0) |  |
| *Language of questionnaire* |  |  | 0.673<sup>d</sup> |
| German | 564 (75.1) | 336 (74.0) |  |
| French | 158 (21.0) | 104 (22.9) |  |
| Italian | 29 (3.9) | 14 (3.1) |  |
| *Migration background<sup>a</sup>* |  |  | **0.027** |
| No | 621 (82.7) | 374 (82.4) |  |
| Yes | 92 (12.3) | 80 (17.6) |  |
| Missing | 38 (5.1) | 0 (0.0) |  |
| *Educational achievement<sup>a</sup>* |  |  | 0.731 |
| Compulsory schooling | 80 (10.7) | 40 (8.8) |  |
| Vocational training | 376 (50.1) | 234 (51.5) |  |
| Upper secondary<br>education | 130 (17.3) | 75 (16.5) |  |
| University education | 115 (15.3) | 73 (16.1) |  |
| Missing | 50 (6.7) | 32 (7.1) |  |
| *Employment status<sup>a</sup>* |  |  | **0.044** |
| Employed | 419 (55.8) | 219 (48.2) |  |
| Unemployed | 64 (8.5) | 45 (9.9) |  |
| Retired | 250 (33.3) | 177 (39.0) |  |
| Missing | 18 (2.4) | 13 (2.9) |  |
| *Partnership<sup>a</sup>* |  |  | **0.007** |
| Yes | 659 (87.7) | 375 (82.6) |  |
| No | 74 (9.9) | 68 (15.0) |  |
| Missing | 18 (2.4) | 11 (2.4) |  |
| *Chronic condition or health problem<sup>a</sup>* |  |  | **0.024** |
| No | 414 (55.1) | 218 (48.0) |  |
| Yes | 330 (43.9) | 228 (50.2) |  |
| Missing | 7 (0.9) | 8 (1.8) |  |

|  | Mean (SD) | Mean (SD) |  |
| --- | --- | --- | --- |
| *Age at study (years)<sup>a</sup>* | 62.1 (6.8) | 62.0 (7.9) | 0.823 |
| **Cancer-related characteristics** |  |  |  |
|  | n (%) |  |  |
| *Late effects survivor (self-reported)<sup>b</sup>* |  |  | - |
| No | 260 (34.6) | - |  |
| Yes | 181 (24.1) | - |  |
| Not assessed | 297 (39.5) | - |  |

*Note:* CCS, childhood cancer survivor; SD, standard deviation
<sup>a</sup> Missing values; percentages are based on the total number of participants
<sup>b</sup> Information only available for group of parents of CCS who were informed about the study aim (n = 454, 60.5% of participating parents of CCS)
<sup>c</sup> p value for comparison of the two samples (Chi-squared test for categorical variables, t-test for continuous variables); significant p values are indicated in bold.
<sup>d</sup> p value for Chi-squared test German vs. French or Italian

For aim 1), we carried out linear regression models using a multilevel approach to assess differences in mean HRQOL (summary measures and health domain scales) between parents of CCS and comparison parents. We included random intercepts, constant slopes, and survivor (parents of CCS) or household (comparison parents) as the group variable to account for family/household clustering. To evaluate the differences between mothers of CCS and comparison mothers, and separately between fathers of CCS and comparison fathers, we calculated linear regression models. We adjusted all models for the adjustment variables. Positive differences indicate better HRQOL and negative differences poorer HRQOL in parents of CCS compared to comparison parents. P values for differences were derived from the linear regression models. For aim 2), we run univariable and multivariable linear regression models using the same multilevel approach. Characteristics associated with HRQOL in the univariable regression model (p<0.05) were included in the multivariable regression models. For the multivariable regression analysis, the variable “mentioning of study aim” was excluded due to the correlation with the assessment of late effects.

## Results

### Sample characteristics

A total of 787 parents of long-term CCS participated in the questionnaire survey (response rate=53.7%). Sociodemographic and cancer-related characteristics of CCS from participating and non-participating parents were comparable (Table S1 in Supplementary Material). Parents of CCS contacted with the cancer-related study aim were more likely to participate compared to parents of CCS with the well-being study aim (p<0.001). Of all participating parents, 751 parents (59% mothers; mean age=62.1 years, SD=6.8 years; Table 1) of 493 CCS completed the SF-36v2 questionnaire and were included in the analyses. Of 258 CCS, both parents participated (n=516) and of 235 CCS, one parent participated. Among CCS with one parent participating, 78% were mothers (n=184) and 22% were fathers (n=51). In the Swiss general population sample, SF-36v2 data were available for 1209 persons of whom 454 had at least one child aged ≥20 years at the time of the study. Thus, a total of 454 comparison parents from 342 households were included in the current analyses as comparison parents (58% mothers; mean age=62.0 years, SD=7.9 years; Table 1).

### Comparison of HRQOL between parents of CCS and comparison parents

Mean scores of the health domain scales and the summary measures for physical and mental HRQOL for the total sample of CCS parents and separately for mothers and fathers are displayed in Table 2. Parents of CCS reported similar HRQOL as comparison parents for physical and mental HRQOL and in all individual health domain scales (Figure 1, Table S2). Mothers of CCS and of the general population had similar HRQOL, except for physical functioning, which was higher in mothers of CCS (p=0.038) than in comparison mothers. Fathers of CCS did not differ from comparison fathers in physical or mental HRQOL or any HRQOL domain (Figure 1, Table S2).

**Figure 1.**
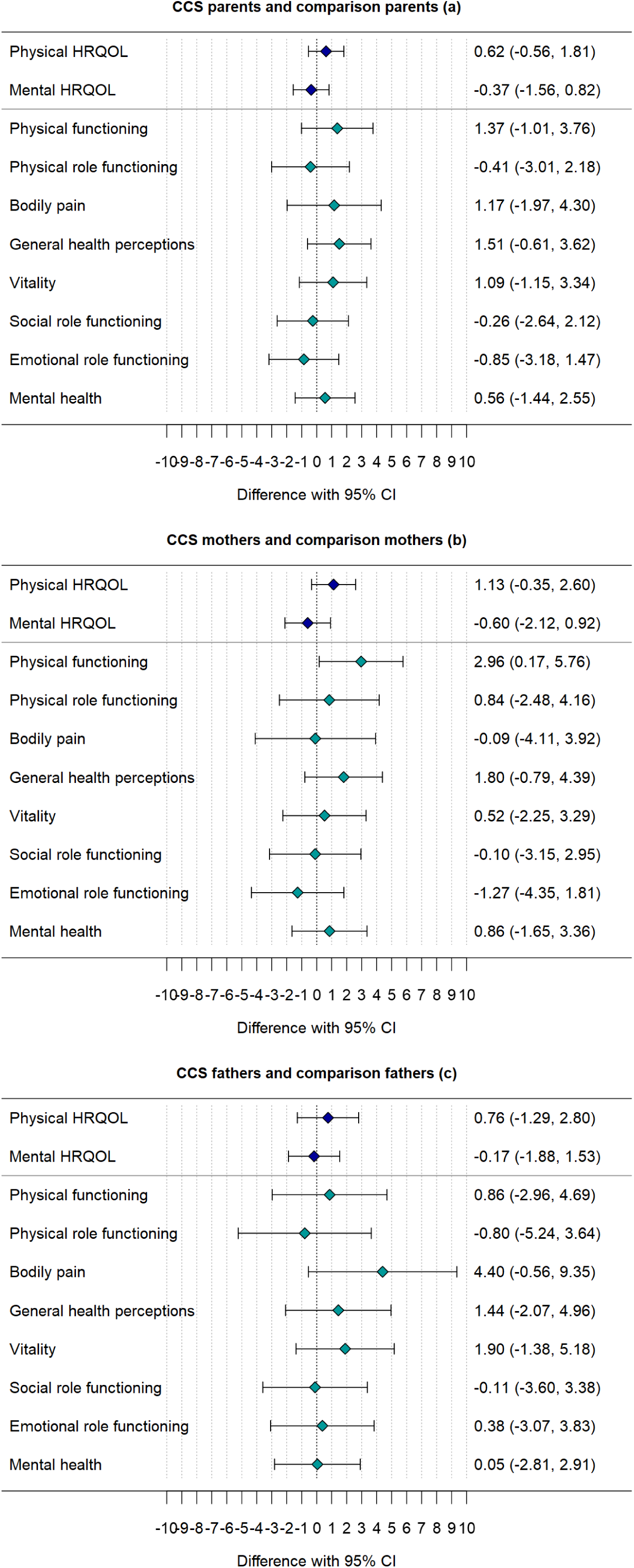
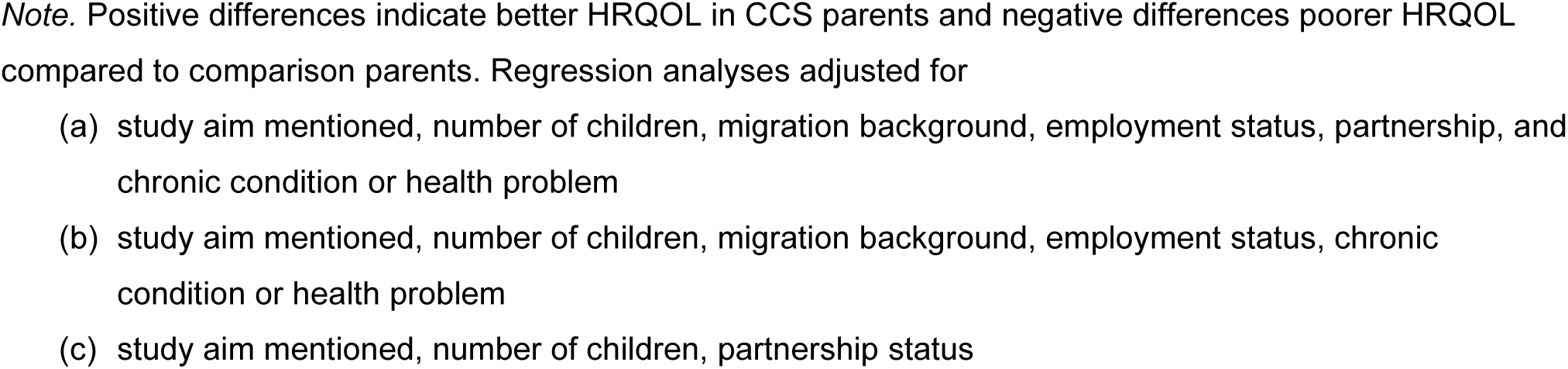
Comparison of HRQOL between parents of childhood cancer survivors (CCS parents; n = 751) and parents from the general population (comparison parents; n = 454) and separately for mothers and fathers for the physical and mental HRQOL summary measures and the health domain scales.

**Table 2.** Mean with 95% confidence interval (CI) in CCS parents (n = 751) and mothers and fathers separately for the health domain scales and the summary measures for physical and mental HRQOL.

|  | Parents of CCS |  | CCS mothers |  | CCS fathers |  |
| --- | --- | --- | --- | --- | --- | --- |
|  | Mean | 95% CI mean | Mean | 95% CI mean | Mean | 95% CI mean |
| **Scale** |  |  |  |  |  |  |
| Physical functioning | 88.87 | (87.62, 90.13) | 88.96 | (87.47, 90.45) | 88.76 | (86.57, 90.94) |
| Physical role functioning | 83.67 | (82.14, 85.20) | 83.24 | (81.35, 85.12) | 84.28 | (81.73, 86.84) |
| Bodily pain | 70.67 | (68.80, 72.54) | 67.77 | (65.32, 70.22) | 74.82 | (71.97, 77.67) |
| General health perceptions | 76.86 | (75.59, 78.13) | 77.27 | (75.70, 78.84) | 76.28 | (74.16, 78.41) |
| Vitality | 68.11 | (66.88, 69.34) | 66.36 | (64.74, 67.97) | 70.61 | (68.74, 72.48) |
| Social role functioning | 88.53 | (87.28, 89.79) | 87.81 | (86.18, 89.44) | 89.56 | (87.59, 91.53) |
| Emotional role functioning | 88.72 | (87.42, 90.01) | 87.57 | (85.87, 89.26) | 90.36 | (88.36, 92.36) |
| Mental health | 78.32 | (77.28, 79.36) | 77.3 | (75.96, 78.65) | 79.78 | (78.15, 81.40) |
| **Summary measure** |  |  |  |  |  |  |
| Physical HRQOL | 47.91 | (47.19, 48.62) | 47.74 | (46.85, 48.62) | 48.14 | (46.96, 49.32) |
| Mental HRQOL | 52.54 | (51.90, 53.18) | 51.89 | (51.04, 52.74) | 53.48 | (52.51, 54.44) |

*Note:* CCS, childhood cancer survivor; CI, confidence interval; HRQOL, health-related quality of life.

### Determinants for lower HRQOL

Results for the univariable regression analyses can be found in Table S3. The multivariable regression analysis showed that low educational attainment, being from the French or Italian part of Switzerland, and having chronic health conditions were determinants for low physical HRQOL in parents of CCS (Figure 2, Table S4). Being a mother, having a migration background, being from the French or Italian part of Switzerland, and having chronic health conditions were determinants for low mental HRQOL (Figure 3, Table S4) in CCS parents.

**Figure 2.**
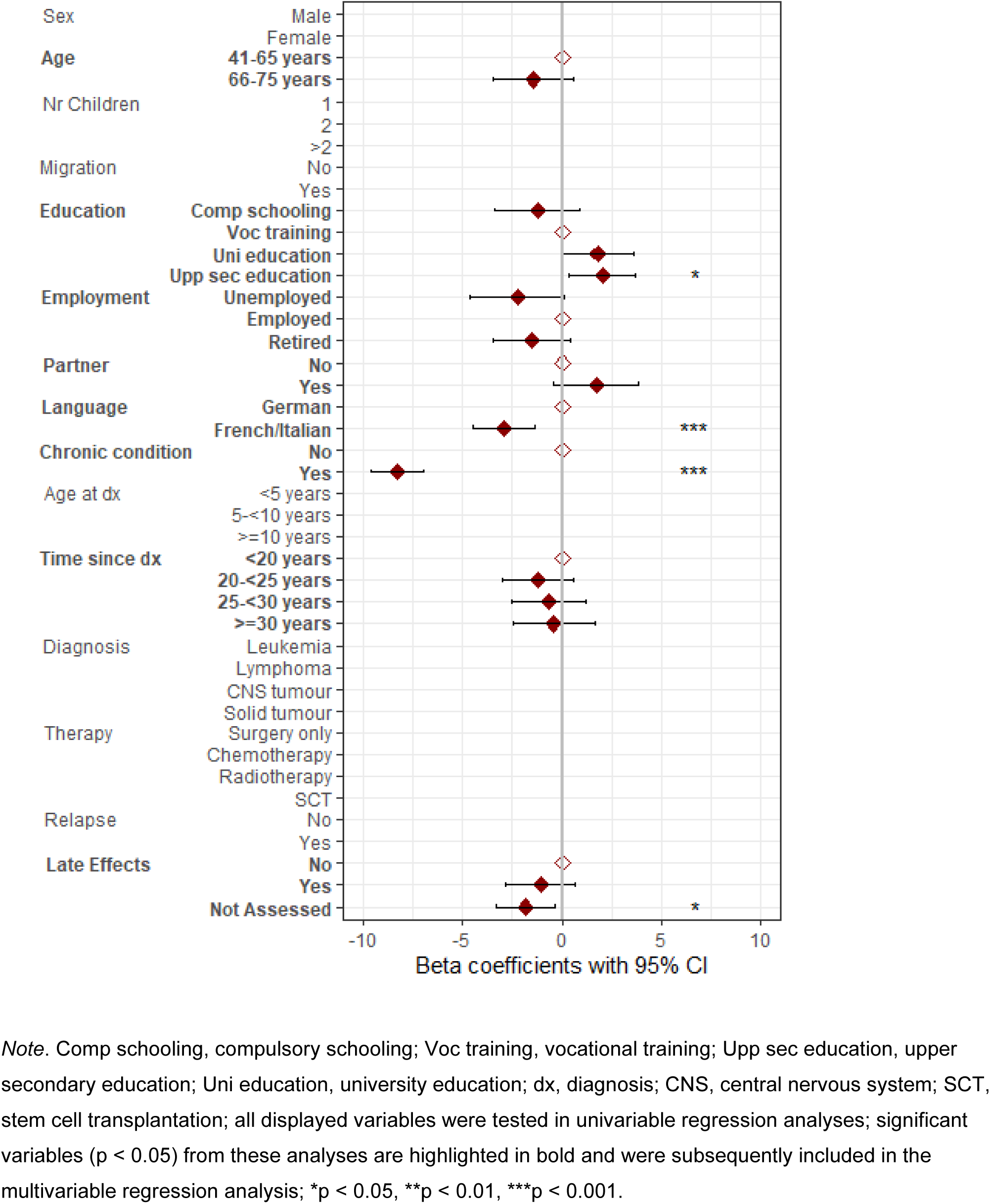
Multilevel multivariable linear regression analysis for physical health-related quality of life (HRQOL) including 657 parents of 453 childhood cancer survivors (parents of CCS)

**Figure 3.**
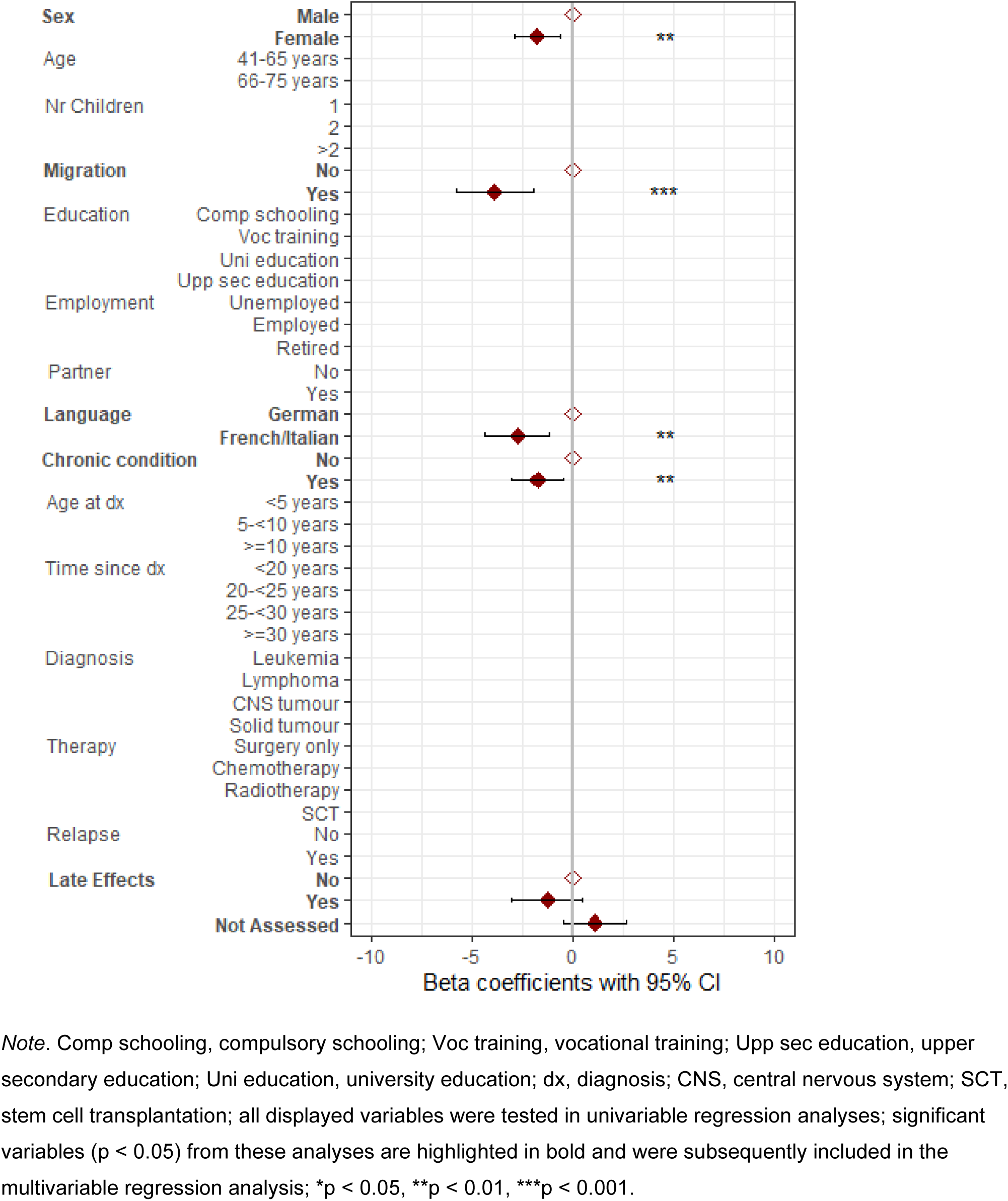
Multilevel multivariable linear regression analysis for mental health-related quality of life (HRQOL) including 695 parents of 471 childhood cancer survivors (parents of CCS).

## Discussion

On average 24 years after their child’s cancer diagnosis, parents of CCS reported physical and mental HRQOL comparable to that of parents in the general population. Mothers and fathers of CCS showed similar HRQOL compared to their respective comparison mothers and fathers, except for higher HRQOL in the domain of physical functioning in mothers of CCS. Several sociodemographic characteristics were associated with low HRQOL, whereas none of the cancer-related characteristics were associated with HRQOL in parents of CCS.

Regarding our first aim, we found that parents of CCS reported similar levels of physical and mental HRQOL to comparison parents. This finding aligns with a previous study, which showed that the overall HRQOL of CCS parents did not differ from normative data 10 years post-diagnosis [8]. Similarly, a Dutch study reported comparable or slightly better HRQOL in parents of young adult CCS compared to a reference sample 21 years after diagnosis [9], however, a French study involving parents of long-term CCS found differing results [7]; while physical HRQOL was lower compared to a reference population, mental HRQOL was higher 7 years after diagnosis.

Furthermore, we found that HRQOL across all health domains was similar in parents of CCS and parents from the general population. This contrasts with a previous study showing differences in HRQOL in all domains except bodily pain [8]. Specifically, a higher proportion of parents from the general population reported problems in the domains of mobility, self-care, and usual activities, whereas a higher proportion of CCS parents reported problems in the domain of mental health. Comparing mothers and fathers separately revealed that mothers of CCS had similar HRQOL levels in all domains except physical functioning, where they fared better than comparison mothers. Fathers of CCS did not differ from comparison fathers in any HRQOL domain. A previous study found that mothers of CCS had higher HRQOL in all domains except sleep and cognitive functioning compared to a female reference sample. Additionally, CCS fathers had significantly higher HRQOL in social functioning and aggressive emotions compared to reference males [9].

Differences in methodological approaches may explain the varying results when comparing HRQOL in our sample of parents to those in previous studies. Our sample of CCS parents differed from the comparison parent sample in several sociodemographic characteristics, which we adjusted for in the analyses. Previous studies used reference samples of adults in general, not specifically parents, and did not compare or account for differing sociodemographic characteristics. Another possibility is that CCS parents in the other studies received different psychological and social support during their child’s cancer journey. Access to support groups, mental health services, and community resources can influence how parents cope with their child’s illness and recovery, thereby impacting HRQOL [26].

Addressing aim 2, we found that none of the cancer-related characteristics were associated with mental or physical HRQOL in parents of CCS. This finding is consistent with a previous study [8]. However, other research on cancer-related determinants of HRQOL in parents of long-term CCS does not present a consistent picture. For instance, a French study found that a longer time since diagnosis was associated with higher psychological QOL [7]. This supports findings from longitudinal studies [3, 4] suggesting that HRQOL gradually improves with time since diagnosis. However, given the range of time since diagnosis in our study (7-40 years), this factor might be less influential, as even the shortest duration since diagnosis is still relatively long. A Dutch study found that cancer recurrence was a significant predictor of higher HRQOL in the social and emotional domains. [9]. Another study found lower HRQOL was significantly predicted by certain types of diagnosis and higher treatment intensity, but only in fathers [5]. Since we did not examine the associations with cancer-related characteristics separately for mothers and fathers, a direct comparison with our findings is not possible.

Mothers of CSS in our study exhibited lower mental HRQOL compared to fathers, but physical HRQOL was similar. This finding aligns with previous studies involving parents of CCS, showing that mothers have significantly lower mental [6] and psychological [7] HRQOL compared to fathers. In the general population (N=1209), men also reported higher mental HRQOL than women in Switzerland [13], Germany [27] and in the US [28]. Thus, lower mental HRQOL observed in mothers of CCS compared to fathers of CCS in our study is not necessarily related to their child’s illness. Lower mental HRQOL in women has been linked to factors such as less social support [29] and to unfavorable socio-economic characteristics such as lower income [30]. Further exploration and understanding of the gender-specific causes of women’s lower mental HRQOL, both in mothers of CCS and in the general population, are critical.

Additionally, we found that having a chronic health condition was associated with both lower physical and mental HRQOL. The same association was found in a large sample representative of the Swiss population [13] and in a review involving adults with chronic conditions [31]. It is therefore likely that this association is independent of the child’s cancer. The same assumption likely applies to the other determinants of lower HRQOL found in this study. Specifically, being from the French or Italian parts of Switzerland was associated with lower physical and mental HRQOL. Lower education was associated with lower physical HRQOL, while having a migration background was associated with lower mental HRQOL. Similar results were found in the Swiss general population [13]. These findings suggest that our results in the sample of CCS parents may not be related to childhood cancer.

A strength of this study is its relatively large sample sizes and large proportion of participating fathers, which enhance the reliability and generalizability of the findings. Another notable strength is the comprehensive comparison framework, which includes both comparisons between parents of CCS and the general population, as well as between mothers and fathers separately. This approach offers a thorough examination of HRQOL. There are limitations that need to be considered when interpreting the findings. The reliance on self-reported data for assessing HRQOL may have introduced social desirability or recall bias. Nevertheless, the SF-36v2 is a well-validated and widely used instrument for assessing HRQOL. The study was conducted in Switzerland, which may limit the generalizability of the findings to other cultural or geographic contexts, as the sociodemographic characteristics may not be representative of more diverse populations.

In conclusion, this study provides valuable insights into the long-term HRQOL of CCS parents in Switzerland, a population that has been understudied so far. Our findings present a positive picture of parental HRQOL, on average 24 years after their children’s cancer diagnosis, showing that CCS parents have comparable physical and mental HRQOL to parents from the general population. It appears that parents of CCS have recovered from the difficult period during the cancer illness and have adapted well. Differences between subgroups of CCS parents based on sociodemographic characteristics may be explained by factors independent of their child’s cancer. These findings indicate that while supportive care should concentrate on the short-term period following a cancer diagnosis and extend into the subsequent years, the influence of cancer-related factors on HRQOL seems to diminish long after the diagnosis. Therefore, the focus should shift towards supporting parents who are, similar to the general population, at risk for poor HRQOL, such as mothers, parents with migrant background, or parents with educational disadvantages.

## Acknowledgments

We thank all study participants for participating in our surveys, the data managers of the Swiss Paediatric Oncology Group, and the Childhood Cancer Registry (ChCR) of the Institute of Social and Preventive Medicine (ISPM) at the University of Bern, Switzerland.

## Statements and Declarations

### Author contributions

Open access funding provided by University of Lucerne. GM initiated the study. JB, LM, FGP, FN, GM, and KR designed and implemented the study. JB, LM, EH, FGP, FN, GM, and KR collected the data. KR and SK conducted the data analysis. KR and SK were the main contributors to the manuscript. GM secured resources and funding. All authors provided feedback and approved the final version of the manuscript.

### Ethical approval

We received ethical approval from the Ethics Committee of Northwest and Central Switzerland (EKNZ 2015-075; 26 March 2015). The survey was conducted in line with the ethical principles of the WMA Declaration of Helsinki.

### Consent to participate

Written informed consent was obtained from all individual participants included in the study.

### Data availability

Requests for data from the Swiss Childhood Cancer Survivor Study–Parents (SCCSS-Parents) should be communicated to the corresponding author. Individual-level, fully anonymized, sensitive data can only be made available to researchers who fulfill the respective legal requirements. Requests for data from the Childhood Cancer Registry must be directed to the Childhood Cancer Registry of Switzerland (https://www.childhoodcancerregistry.ch/).

