## Supplementary Material for "Health-related quality of life in parents of long-term childhood cancer survivors: a report from the Swiss Childhood Cancer Survivor Study - Parents"

**Table S1**

*Comparison of survivor characteristics with participating (n = 493) and non-participating (n = 734) parents.*

| | **Survivors with participating parents (n = 493)** | **Survivors with non-participating parents (n = 734)** | |
|---|---|---|---|
| | **n (%)** | **n (%)** | **p value [b]** |
| *Study aim* | | | **<0.001** |
| Cancer-related study aim | 296 (60.0) | 310 (42.2) | |
| Well-being study aim | 197 (40.0) | 424 (57.8) | |
| **Socio-demographic characteristics** | | | |
| *Survivors' sex* | | | 0.636 |
| Male | 272 (55.2) | 415 (56.5) | |
| Female | 221 (44.8) | 319 (43.5) | |
| *Survivors' age at study* | | | 0.065 |
| <30 years | 209 (42.4) | 298 (40.6) | |
| 30-<40 years | 218 (44.2) | 301 (41.0) | |
| >=40 years | 66 (13.4) | 135 (18.4) | |
| | **Mean (SD)** | **Mean (SD)** | **p value [b]** |
| *Survivors' age at study (years)* | 32 (6.2) | 33 (6.8) | 0.113 |
| **Cancer-related characteristics** | | | |
| | **n (%)** | **n (%)** | **p value [b]** |
| *Age at diagnosis* | | | 0.400 |
| <5 years | 188 (38.1) | 270 (36.8) | |
| 5-<10 years | 143 (29.0) | 196 (26.7) | |
| >=10 years | 162 (32.9) | 268 (36.5) | |
| *Time since diagnosis* | | | 0.232 |
| <20 years | 140 (28.4) | 190 (25.9) | |
| 20-<25 years | 125 (25.4) | 210 (28.6) | |
| 25-<30 years | 121 (24.5) | 155 (21.1) | |
| >=30 years | 107 (21.7) | 179 (24.4) | |
| *Diagnosis (ICCC-3)* | | | 0.803 |
| Leukemia | 167 (33.9) | 248 (33.8) | |
| Lymphoma | 84 (17.0) | 133 (18.1) | |
| CNS tumor | 68 (13.8) | 109 (14.9) | |
| Neuroblastoma | 18 (3.7) | 37 (5.0) | |

| | | | |
|---|---|---|---|
| Retinoblastoma | 14 (2.8) | 13 (1.8) | |
| Renal tumor | 33 (6.7) | 50 (6.8) | |
| Hepatic tumor | 6 (1.2) | 4 (0.5) | |
| Bone tumor | 29 (5.9) | 33 (4.5) | |
| Soft tissue sarcoma | 33 (6.7) | 46 (6.3) | |
| Germ cell tumor | 17 (3.4) | 27 (3.7) | |
| LCH | 24 (4.9) | 34 (4.6) | |
| *Therapy* [a] | | | 0.178 |
| Surgery only | 64 (13.0) | 90 (12.3) | |
| *Chemotherapy* | 263 (53.3) | 358 (48.8) | |
| Radiotherapy | 139 (28.2) | 249 (33.9) | |
| SCT | 26 (5.3) | 32 (4.4) | |
| *Relapse* | | | 0.402 |
| No | 432 (87.6) | 631 (86.0) | |
| Yes | 61 (12.4) | 103 (14.0) | |
| | **Mean (SD)** | **Mean (SD)** | **p value** [b] |
| *Age at diagnosis (years)* | 7 (4.5) | 7 (4.6) | 0.578 |
| *Time since diagnosis (years)* | 24 (6.8) | 24 (7.0) | 0.292 |

*Note*. CNS, central nervous system; LCH, Langerhans cell histiocytosis; SCT, stem cell transplantation; SD, standard deviation

[a] Missing values; percentages are based on the total number of participants

[b] p value calculated from Chi-square test statistics (categorical variables) or t test statistics (continuous variables) comparing survivors of participating and non-participating parents.

**Table S2**

*Comparison of HRQOL between parents of childhood cancer survivors (CCS parents; n = 751) and parents from the general population (comparison parents; n = 454) and separately for mothers and fathers for the physical and mental HRQOL summary measures and the health domain scales and corresponding p values.*

| | **Difference between CCS parents (n = 751) and comparison parents (n = 454) [a]** | | |
|---|---|---|---|
| | **Difference** | **95% CI difference** | **p value** |
| **Scale** | | | |
| Physical functioning | 1.37 | (-1.01, 3.76) | 0.259 |
| Physical role functioning | -0.41 | (-3.01, 2.18) | 0.755 |
| Bodily pain | 1.17 | (-1.97, 4.30) | 0.465 |
| General health perceptions | 1.51 | (-0.61, 3.62) | 0.163 |
| Vitality | 1.09 | (-1.15, 3.34) | 0.340 |
| Social role functioning | -0.26 | (-2.64, 2.12) | 0.831 |
| Emotional role functioning | -0.85 | (-3.18, 1.47) | 0.472 |
| Mental health | 0.56 | (-1.44, 2.55) | 0.583 |
| **Summary measure** | | | |
| Physical HRQOL | 0.62 | (-0.56, 1.81) | 0.300 |
| Mental HRQOL | -0.37 | (-1.56, 0.82) | 0.545 |
| | **Difference between CCS mothers (n = 442) and comparison mothers (n = 263) [b]** | | |
| | **Difference** | **95% CI difference** | **p value** |
| **Scale** | | | |
| Physical functioning | 2.96 | (0.17, 5.76) | **0.038** |
| Physical role functioning | 0.84 | (-2.48, 4.16) | 0.620 |
| Bodily pain | -0.09 | (-4.11, 3.92) | 0.964 |
| General health perceptions | 1.80 | (-0.79, 4.39) | 0.173 |
| Vitality | 0.52 | (-2.25, 3.29) | 0.711 |
| Social role functioning | -0.10 | (-3.15, 2.95) | 0.950 |
| Emotional role functioning | -1.27 | (-4.35, 1.81) | 0.418 |
| Mental health | 0.86 | (-1.65, 3.36) | 0.503 |
| **Summary measure** | | | |
| Physical HRQOL | 1.13 | (-0.35, 2.60) | 0.134 |
| Mental HRQOL | -0.60 | (-2.12, 0.92) | 0.437 |
| | **Difference between CCS fathers (n = 309) and comparison fathers (n = 191) [c]** | | |
| | **Difference** | **95% CI difference** | **p value** |
| **Scale** | | | |
| Physical functioning | 0.86 | (-2.96, 4.69) | 0.658 |
| Physical role functioning | -0.80 | (-5.24, 3.64) | 0.724 |
| Bodily pain | 4.40 | (-0.56, 9.35) | 0.082 |
| General health perceptions | 1.44 | (-2.07, 4.96) | 0.420 |
| Vitality | 1.90 | (-1.38, 5.18) | 0.255 |

| | | | |
|---|---|---|---|
| Social role functioning | -0.11 | (-3.60, 3.38) | 0.951 |
| Emotional role functioning | 0.38 | (-3.07, 3.83) | 0.829 |
| Mental health | 0.05 | (-2.81, 2.91) | 0.974 |
| **Summary measure** | | | |
| Physical HRQOL | 0.76 | (-1.29, 2.80) | 0.467 |
| Mental HRQOL | -0.17 | (-1.88, 1.53) | 0.841 |

*Note.* CCS, childhood cancer survivor; CI, confidence interval; HRQOL, health-related quality of life; difference with 95% confidence interval (CI) comparing CCS parents and comparison parents; positive differences indicate better HRQOL in CCS parents and negative differences poorer HRQOL compared to comparison parents; p value for the adjusted difference between the two samples; p values <0.05 are indicated with bold font.

Regression analyses adjusted for

[a] study aim mentioned, number of children, migration background, employment status, partnership, and chronic condition or health problem

[b] study aim mentioned, number of children, migration background, employment status, and chronic condition or health problem

[c] study aim mentioned, number of children, and partnership status

**Table S3**

*Multilevel univariable linear regression analysis for physical and mental health-related quality of life (HRQOL) including parents of childhood cancer survivors (CCS parents).*

| | Physical HRQOL (PCS) | | | Mental HRQOL (MCS) | | |
|---|---|---|---|---|---|---|
| ***Characteristics*** | **Coeff** | **95% CI** | **p value [b]** | **Coeff** | **95% CI** | **p value [b]** |
| **Study aim mentioned** | | | 0.253 | | | **0.036** |
| No | Ref | | | Ref | | |
| Yes | 0.89 | (-0.63, 2.40) | | -1.54 | (-2.99, -0.10) | |
| ***Socio-demographic characteristics*** | | | | | | |
| **Sex** | | | 0.681 | | | **0.006** |
| Fathers | Ref | | | Ref | | |
| Mothers | -0.29 | (-1.65, 1.08) | | -1.55 | (-2.65, -0.44) | |
| **Age [a]** | | | **<0.001** | | | 0.476 |
| 41-65 years | Ref | | | Ref | | |
| 66-75 years | -4.17 | (-5.70, -2.63) | | 0.53 | (-0.92, 1.98) | |
| **Number of children [a]** | | | 0.246 | | | 0.654 |
| 1 child | Ref | | | Ref | | |
| 2 children | 2.03 | (-2.20, 6.25) | | -1.82 | (-5.70, 2.06) | |
| >2 children | 0.81 | (-3.41, 5.03) | | -1.65 | (-5.54, 2.24) | |
| **Migration background [a]** | | | 0.430 | | | **<0.001** |
| No | Ref | | | Ref | | |
| Yes | -0.87 | (-3.03, 1.29) | | -3.87 | (-5.82, -1.91) | |
| **Education [a]** | | | **0.046** | | | 0.338 |
| Compulsory schooling | -0.53 | (-3.87, 0.82) | | -0.58 | (-2.73, 1.58) | |
| Vocational training | Ref | | | Ref | | |
| Upper secondary education | 1.86 | (-0.04, 3.77) | | 0.78 | (-0.93, 2.50) | |
| University education | 1.38 | (-0.67, 3.43) | | 1.43 | (-0.47, 3.33) | |
| **Employment [a]** | | | **<0.001** | | | 0.171 |
| Employed | Ref | | | Ref | | |
| Unemployed | -3.48 | (-6.01, -0.92) | | -2.12 | (-4.38, 0.14) | |

| | | | | | | |
|---|---|---|---|---|---|---|
| Retired | -3.76 | (-5.29, -2.23) | | -0.06 | (-1.50, 1.37) | |
| **Partnership [a]** | | | **0.003** | | | 0.108 |
| Yes | Ref | | | Ref | | |
| No | -3.59 | (6.00, -1.19) | | -1.81 | (-4.01, 0.40) | |
| **Questionnaire language** | | | **<0.001** | | | **<0.001** |
| German | Ref | | | Ref | | |
| French or Italian | -3.32 | (-5.00, -1.63) | | -3.48 | (-5.07, -1.89) | |
| **Chronic condition or health problem [a]** | | | **<0.001** | | | **0.011** |
| No | Ref | | | Ref | | |
| Yes | -8.61 | (-9.91, -7.32) | | -1.61 | (-2.86, -0.36) | |
| ***Cancer-related characteristics*** | | | | | | |
| **Age at diagnosis** | | | 0.093 | | | 0.543 |
| <5 years | Ref | | | Ref | | |
| 5-<10 years | -1.95 | (-3.75, -0.159 | | -0.11 | (-1.84, 1.62) | |
| >= 10 years | -1.24 | (-3.01, 0.53) | | -0.90 | (-2.59, 0.79) | |
| **Time since diagnosis** | | | **0.002** | | | 0.921 |
| <20 years | Ref | | | Ref | | |
| 20-<25 years | -1.80 | (-3.78, 0.19) | | -0.37 | (-2.30, 1.56) | |
| 25-<30 years | -2.72 | (-4.74, -0.71) | | -0.44 | (-2.39, 1.51) | |
| >=30 years | -3.92 | (-6.04, -1.80) | | 0.19 | (-1.85, 2.23) | |
| **Cancer diagnosis** | | | 0.147 | | | 0.808 |
| Leukemia | Ref | | | Ref | | |
| Lymphoma | -2.21 | (-4.24, -0.17) | | -0.97 | (-2.91, 0.97) | |
| CNS tumour | -0.98 | (-3.35, 1.40) | | -0.45 | (-2.72, 1.81) | |
| Solid tumour | -0.13 | (-1.98, 1.72) | | -0.49 | (-2.25, 1.28) | |
| **Therapy** | | | 0.626 | | | 0.086 |
| Surgery only | Ref | | | Ref | | |
| Chemotherapy | 0.45 | (-1.82, 2.71) | | 1.69 | (-0.46, 3.84) | |
| Radiotherapy | -0.49 | (-2.97, 1.99) | | -0.24 | (-2.59, 2.11) | |
| SCT | 1.49 | (-2.38, 5.35) | | 1.91 | (-1.75, 5.56) | |
| **Relapse** | | | 0.506 | | | 0.470 |
| No | Ref | | | Ref | | |
| Yes | 0.76 | (-1.49, 3.01) | | -0.79 | (-2.94, 1.35) | |
| **Late effects survivor (self-reported) [b]** | | | **0.026** | | | **0.006** |
| No | Ref | | | Ref | | |
| Yes | -2.23 | (-4.13, -0.33) | | -2.09 | (-3.83, -0.35) | |

| | | | | |
|---|---|---|---|---|
| Not assessed | -2.01 | (-3.72, 0.31) | 0.69 | (-0.92, 2.31) |

*Note*. HRQOL, health-related quality of life; PCS, physical component summary; MCS, mental component summary; n, number; Coeff, coefficient; CI, confidence interval; Ref, reference category

p values <0.05 are indicated with bold font.

[a] Missing values

[b] p values calculated from Wald tests

**Table S4**

*Multilevel multivariable linear regression analyses for physical and mental health-related quality of life (HRQOL) including parents of childhood cancer survivors (CCS parents).*

| | **Physical HRQOL (PCS) (n parents = 657, n groups = 453)** | | | **Mental HRQOL (MCS) (n parents = 695, n groups = 471)** | | |
|---|---|---|---|---|---|---|
| **Characteristics** | **Coeff** | **95% CI** | **p value [c]** | **Coeff** | **95% CI** | **p value [c]** |
| **Study aim mentioned [a]** | | | - | | | - |
| No | - | - | | - | - | |
| Yes | - | - | | - | - | |
| **Sex** | | | - | | | **0.002** |
| Fathers | - | - | | Ref | | |
| Mothers | - | - | | -1.80 | (-2.93, -0.66) | |
| **Age [b]** | | | 0.160 | | | - |
| 41-65 years | Ref | | | - | - | |
| 66-75 years | -1.43 | (-3.43, 0.56) | | - | - | |
| **Migration background [b]** | | | - | | | **0.000** |
| No | - | - | | Ref | | |
| Yes | - | - | | -3.88 | (-5.79, -1.97) | |
| **Education [b]** | | | **0.012** | | | - |
| Compulsory schooling | -1.23 | (-3.38, 0.92) | | - | - | |
| Vocational training | Ref | | | - | - | |
| Upper secondary education | 2.01 | (0.32, 3.71) | | - | - | |
| University education | 1.79 | (-0.04, 3.62) | | - | - | |
| **Employment [b]** | | | **0.000** | | | - |
| Employed | Ref | | | - | - | |
| Unemployed | -2.23 | (-4.57, 0.12) | | - | - | |
| Retired | -1.55 | (-3.49, 0.39) | | - | - | |
| **Partnership [b]** | | | **0.116** | | | - |
| Yes | Ref | | | - | | |
| No | -1.72 | (-3.87, 0.43) | | - | - | |
| **Questionnaire language** | | | **0.000** | | | **0.001** |

| | | | | | | |
|---|---|---|---|---|---|---|
| German | Ref | | | Ref | | |
| French or Italian | -2.91 | (-4.44, -1.38) | | -2.75 | (-4.36, -1.15) | |
| **Chronic condition or health problem [b]** | | | **0.000** | | | **0.006** |
| No | Ref | | | Ref | | |
| Yes | -8.26 | (-9.56, -6.96) | | -1.76 | (-3.02, -0.51) | |
| **Time since diagnosis** | | | **0.585** | | | |
| <20 years | Ref | | | | | |
| 20-<25 years | -1.21 | (-2.96, 0.54) | | | | |
| 25-<30 years | -0.68 | (-2.56, 1.19) | | | | |
| >=30 years | -0.41 | (-2.48, 1.67) | | | | |
| **Late effects survivor (self-reported) [b]** | | | **0.057** | | | **0.029** |
| No | Ref | | | Ref | | |
| Yes | -1.09 | (-2.83, 0.64) | | -1.27 | (-3.03, 0.48) | |
| Not assessed | -1.81 | (-3.30, -0.33) | | 1.09 | (-0.49, 2.67) | |

*Note*. HRQOL, health-related quality of life; PCS, physical component summary; MCS, mental component summary; n, number; Coeff, coefficient; CI, confidence interval; Ref, reference category

p values <0.05 are indicated with bold font.

[a] Excluded in multivariable regression analysis due to its correlation with late effects

[b] Missing values

[c] p values calculated from Wald tests
